# Evaluation of the Bangkok Health Research and Ethics Interest Group: reflecting on the experiences of group members, researchers and facilitators participating in an urban community advisory board in Thailand

**DOI:** 10.1101/2025.06.24.25330175

**Authors:** Anne Osterrieder, Supanat Ruangkajorn, Bhensri Naemiratch, Tassawan Poomchaichote, Supa-at Asarath, Kanpong Boonthaworn, Phaik Yeong Cheah

## Abstract

**Background:** The Mahidol Oxford Tropical Medicine Research Unit (MORU), headquartered in Bangkok, conducts research on tropical medicine and global health. MORU works closely with a network of community advisory boards (CABs), with members drawn from communities served by its research sites. In 2019, we set up the Bangkok Health Research and Ethics Interest Group (HREIG). HREIG is an urban CAB composed of Bangkok residents from all walks of life who have an interest in health research, but do not have a professional health research background. HREIG advises on clinical research and broader societal issues, complementing MORU’s rural CABs in areas such as the Thai-Myanmar border and Cambodia. In 2022, the group had 14 members aged 22 to 51. Initially, meetings were held in person and in English, to support discussion with international researchers. During COVID-19 restrictions, meetings moved online and adopted Thai as the main language.

**Main body:** To evaluate the group’s first two years, we conducted a mixed-methods study using focus group discussions and anonymous online surveys with group members, researchers and facilitators. We found that members joined to contribute to research and society, and for personal benefits such as learning about health research or meeting new people. While some valued practicing English, it made harder for members to contribute equally, and even those with good conversational English struggled with technical terms. Researchers reported that engaging with the group helped improve their communication skills, and that they valued hearing lay perspectives. Facilitators strengthened their skills in areas such as facilitation, online collaboration, and explaining research in lay terms. As HREIG is not tied to a specific study, its contributions have ranged from improving malaria messaging for the Thai public to testing COVID-19 surveys, or sharing opinions on vaccines and clinical trial participation.

**Conclusion:** Our findings offer an in-depth insight into the experiences of HREIG members, researchers and facilitators. We share the outcomes of the group’s activities, and the lessons learnt from running a CAB in an international research environment. We hope that our findings will help others who would like to set up a CAB in a similar context.

**Plain English summary:** The Mahidol Oxford Tropical Medicine Research Unit (MORU), headquartered in Bangkok, carries out research on tropical medicine and global health. MORU works closely with a network of community advisory boards (CABs). In 2019, we set up the Bangkok Health Research and Ethics Interest Group (HREIG). Its members are Bangkok residents from all walks of life who have an interest in health research, but do not have a professional health research background. HREIG advises on clinical research and broader societal issues, such as data sharing or artificial intelligence. By 2022, the group had 14 members aged 22 to 51. Initially, meetings were held in English and in person; during COVID-19 restrictions, they shifted online and adopted Thai as the main language.

After two years, we asked members, researchers and facilitators for their feedback on the group. HREIG members had joined the group to contribute to research and society, and for personal benefits, such as learning about health research or meeting new people. Although some members enjoyed practicing English, it made harder for members to contribute equally. Researchers had improved their communication skills, and valued hearing from non-scientists. Facilitators had expanded their professional skills as well. The group’s feedback was important for many studies, including improving communications about malaria for the Thai public, testing COVID-19 survey questions or sharing views about vaccines.

Our findings offer an in-depth insight into the experiences of HREIG members, researchers and facilitators. We hope that our findings will help others who would like to set up a CAB in a similar setting.

**Thaiclinicaltrials.org** **registration: TCTR20211012003**

## Background

Community advisory board (CABs) and public advisory groups are platforms that facilitate consultation and collaboration between researchers and members of the communities that are affected by their research.^1–4^ Involving communities at all stages of the research cycle, particularly at the early stages (e.g. in research or protocol development) can help align research with the needs of communities, build relationships and trust with communities involved in or affected by the research, and make research more relevant and ethical.^1^ ^5–8^ This is especially important for health research conducted in low- and mid-income countries (LMICs), which faces additional challenges around avoiding participant exploitation, and coercion, and being sensitive to local contexts, cultures and societal norms.^9^

There is an increasing number of publications about CABs, or public and patient involvement (PPI) groups, as often referred to in high-income countries, but only few published reports of CABs in Southeast Asia and Thailand^4^, where this case study is set. Examples of different CAB ‘models’ from Southeast Asia include: CAB members who live in the same area and advise on different studies conducted in their communities (e.g. the Tak Province Community Ethics Advisory Board or T-CAB comprises migrants and communities living at the Thai-Myanmar border^10–12^); members who share similar demographics (e.g. youth advisory groups^13^ ^14^, Chiang Rai hill tribe communities^9^); or members who share the lived experience of a health condition (e.g. tuberculosis^15^, HIV/AIDS^16^).

The Mahidol-Oxford Tropical Medicine Research Unit (MORU) Tropical Health Network is a large international research network with major research units including in Thailand, Cambodia, Laos and Myanmar, and collaborative clinical research sites in low-resource settings across the world. Its 900 researchers and support staff conduct clinical and public health research studies and clinical trials on infectious diseases (e.g. tuberculosis, malaria, COVID-19), and maternal and child health. MORU runs CABs in Thailand, Cambodia and Laos, all of which serve different purposes.^17^ ^9^ ^14^ Through our CAB network, we aim to get a broad range of views from the different areas where we conduct our research.

Bangkok is Thailand’s capital and its main urban area, with an estimated population of 11,233,900 million people in 2024, making up about 13% of Thailand’s total population.^18^ Compared to other regions in Thailand, Bangkok has a high proportion of people with Thai ethnicity and young people.^19^ ^20^ In 2019, we set up the Bangkok Health Research and Ethics Interest Group (HREIG) to advise researchers running clinical research at the Bangkok site, such as the Malaria Infection Study in Thailand^21^ or large-scale COVID-19 treatment trials^22^, and those working on topics relevant to a wider population, e.g. antimicrobial resistance, mathematical modelling, data sharing in health care.

In this paper, we share findings from our evaluation after two years of running the HREIG, focusing on its operation, impact, and the lessons learned. The objectives of our evaluation were to explore HREIG members’ motivations for joining and their views on the benefits, challenges and impact of the group. We also explored researchers’ experiences of engaging with HREIG and the group’s impact on their work and professional or personal development. Lastly, HREIG facilitators reflected on the facilitation and management of HREIG.

## Main text

### Who was involved with HREIG?

In February 2022, at the time of the evaluation study, the group had 14 members between 22 and 51 years old, 5 men and 9 women. Five members were recruited in July 2019, four 4 in December 2019, and five in December 2020. Recruitment happened via posting an advertisement on a Thai job site, and word of mouth. Selection criteria were that members had to be Thai, 18 years old or older, be fluent in Thai and English, and resident in the Bangkok metropolitan area. They also needed to have an interest in health research/science and be available to attend 4-6 meetings a year. We excluded applicants with postgraduate degrees or professional experience in health research. We also considered demographics like age, gender and career stage, to achieve a diverse group. The group was not set up to be representative of a specific community, in contrast to other CABs, but rather aimed to obtain a range of different perspectives. The group remit and rules were summarised in a “Terms of Reference” document.^23^

Between 2019 and 2022, HREIG had three facilitators, authors AO, SR and PY, who were planning and running meetings. Due to the international background of the facilitator team (German, Thai and Malaysian) and the MORU researcher community, HREIG meetings initially were held in English. Our Thai team members SN, BN, TP and SA helped with translation of English speaker presentations and the live group chat during the meetings. In 2022, we decided to switch the main meeting language to Thai, to make meetings more inclusive for members who felt less comfortable to speak in English.

A range of MORU researchers attended HREIG meetings to receive advice and feedback on their projects, participate in discussion or to purely observe. These included principal investigators or project leads, senior and junior researchers, and PhD students (at MORU, many researchers hold two or more roles, e.g. PhD student and project lead). As mentioned above, some of the researchers were non-Thai speakers and discussed their work in English.

Between August 2019 and January 2020, meetings lasting between 2 and 3 hours were held in the evening from 6-9 pm on the Faculty of Tropical Medicine campus at Mahidol University in Bangkok. Initially, each meeting covered two different research topics, but upon feedback from the group we limited this to one topic per session. Members received a ‘token of appreciation’ of THB 1000 (about £25) per meeting, as compensation for their time and any travel expenses, equivalent to an average day’s salary for a bachelor graduate and about 2.75 times the minimum daily wage.^18^ From January 2020, meetings moved online due to COVID-19 government restrictions on travel and in-person gatherings. For easy and swift communication with members, we used the messenger application ‘LINE’ to inform members of updates and upcoming activities, and schedule session dates via a poll function. Table 1 shows an overview over HREIG meetings from August 2019 to February 2022, including meeting format and outputs/outcomes.

**Table 1.** Overview over the activities of the Bangkok Health Research and Ethics Interest Group (HREIG) between 2019-2022.

| Date | Session topics and activities | Outputs/outcomes for HREIG members and MORU (researchers/facilitators) |
| --- | --- | --- |
| **Aug 2019** | • Orientation meeting: introductions, group rules<br>• Talk: “What is informed consent?”<br>• Hypothetical clinical trial case study | • Increased understanding of research ethics and informed consent<br>• Feedback on what kind of information researchers would need to provide to potential clinical trial participants |
| **Nov 2019** | • Adaptive vaccine trials<br>• Hypothetical case study (outbreak of a deadly virus in Bangkok and potential participation in a randomised clinical vaccine trial) | • Increased understanding of randomised clinical trials and vaccine development<br>• Feedback on members’ perceptions of clinical trials, reasons for participating, and questions/concerns; feedback on what potential participants would want to know before enrolling in the trial |
| **Jan 2020** | • Malaria communications and key messages in Thai<br>• Substandard and falsified (SF) medicines | • Ranked key messages according to importance, informing research communications strategy; feedback on message content and wording<br>• Increased awareness about SF medicines (e.g. recognising, avoiding)<br>• Ideas for SF public engagement activities |
| **March 2020** | • Malaria Infection Study in Thailand (MiST)<sup>24</sup><br>• “Kaa tob taan” (Thai for compensation) in clinical research<br>• COVID-19 outbreak discussion | • Increased understanding of malaria and the MiST study<br>• List of monetary and non-monetary benefits that people would perceive as compensation for participating in a clinical trial (e.g. meals, insurance, medical advice, health checks)<br>• Feedback on perception of risks, safety, and compensation amount |
| **March 2020** | • SEBCOV study (‘Social, ethical and behavioural aspects of COVID-19’)<sup>25</sup><br>• Testing of selected questions for the SEBCOV online survey<sup>26</sup><br>• Discussion: Coping with social distancing | • Increased understanding of COVID-19 and the SEBCOV study<br>• Feedback on improving the content, wording and clarity of survey questions |
| **May 2020** | • Discussion of rumours and misinformation around COVID-19 and treatments<br>• Introduction to the ‘COPCOV’ study (Chloroquine/hydroxychloroquine prevention of coronavirus disease (COVID-19) in the healthcare setting)<sup>27</sup><br>• COPCOV key messages in English and Thai | • List of examples of rumours and misinformation that members had seen<br>• Increased understanding of the COPCOV study and COVID-19<br>• Feedback on clarity and wording of COPCOV key messages used in participant recruitment materials<br>• Feedback on potential concerns that could affect people’s decision to enrol in the trial |
| **Jan 2021** | • Antimicrobial resistance (AMR) | • Increased understanding of antimicrobials and AMR |
|  | • ‘AMR Dialogues’<sup>28</sup> project (participatory dialogues on antimicrobial resistance) | • Piloting planned online engagement activities for the ‘AMR Dialogues’ project<br>• Insight into members’ feelings relating to antimicrobials<br>• Examples of locations and community groups where antimicrobials are used in daily life<br>• List of suggestions for two-way communications with community groups |
| **May 2021** | • Introduction of six new members<br>• Presentation about COVID-19 vaccines<br>• Discussion around vaccine uptake/hesitancy (*note: vaccine not available in Thailand at time of discussion*) | • Increased understanding of COVID-19 vaccines<br>• Insights into perceptions of different vaccine types, preferences, and concerns |
| **Aug 2021** | • Introduction to the “Antibiotic Footprint Calculator”<sup>29</sup> website (at pre-release stage)<br>• Testing of the website | • Increased understanding of antibiotics and antimicrobial resistance<br>• Feedback and comments on website content, graphics, navigation, accessibility |
| **Feb 2022** | • Clinical trials and control groups (“arms”)<br>• PLATCOV study (‘Finding Treatments for COVID-19: A Trial of Antiviral Pharmacodynamics in Early Symptomatic COVID-19’)<sup>30</sup> | • Increased understanding of clinical trials<br>• Feedback on communicating the concept of “control arms” to potential participants/the public<br>• Insights on perceptions of clinical trials and control arms, and information required before joining |
| **Feb 2022** | • Evaluation | • Focus group discussion transcript for qualitative analysis<br>• Survey responses for quantitative and qualitative analysis |

## Methods

### Evaluation of HREIG

To evaluate the HREIG, we used a mixed-methods approach that included conducted focus group discussions (FGDs) with HREIG members, and facilitators, and anonymous, self-administered online surveys distributed to HREIG members and researchers (Table 2).

**Table 2.** Overview over participant groups, inclusion criteria and data collection methods.

| Participant group | Inclusion criteria | Data collection method |
| --- | --- | --- |
| **HREIG members** | Active group member<br>Group member for at least 12 months | Focus group discussion<br>Anonymous, self-administered online survey |
| **Researchers** | Affiliated with MORU or the Faculty for Tropical Medicine, Mahidol University<br>Engaged with HREIG in the last 2 years, either as presenter, discussion participant, or observer | Anonymous, self-administered online survey |
| **Facilitators** | Helped with at least one session (facilitation and translation) in the last two years | Focus group discussion |

Meeting minutes were reviewed to help identify the group’s outcomes and impact. Six HREIG members participated in both the FGDs and survey, 13 researchers completed the survey, and five facilitators participated in the FGD.

### HREIG member focus group discussion and survey

Six HREIG members were invited by email to participate in an FGD. All of them agreed to participate and gave informed consent. They were aged between 22 and 43 years, and their professions were university students, business owner, and professional/administrative employees. The FGD was conducted in English and Thai via Microsoft Teams, lasted 1.5 hours, and was audio recorded (participants were asked to turn off their cameras) with a backup used of digital audio recording device. The FGD topic guide was developed using two frameworks: our HREIG Logic Model, which is a theoretical visualisation of how outputs, outcomes and impact are expected to occur as a result of inputs and activities^31^ (Suppl. Fig. 1); and the ‘UK Standards for Public Involvement’ framework.^32^ ^33^ This framework provides good practice guidelines for public involvement and self-assessment. It consists of six standards (Inclusive Opportunities, Working Together, Support and Learning, Governance, Communications, and Impact) and reflective questions for each standard. From these, we selected the most relevant questions to structure our topic guides.

To give everyone the opportunity to express their opinions honestly and upon individual reflection, we asked members to complete a short, anonymous online survey (via Microsoft Forms), at the start of the FGD. The survey consisted of seven questions and included three Likert-scale questions on reasons for joining the group and their feelings about participation and being a part of the group; and three open-text questions on challenges they experienced as a group member, views on financial compensation for attending meetings, and any additional comments. The survey questions were informed by anonymous feedback gathered at the end of the group induction meeting in August 2019.

### Researcher survey

An anonymous, self-administered online survey was sent to researchers who had engaged with HREIG by email. The survey was set up on the platform JISC Online Surveys.^34^ It consisted of 16 questions in English, which were a mix of single-choice, multiple choice, Likert scale and open text questions. Of the 16 invited researchers, 13 completed the survey. When asked about their level of engagement with HREIG, five respondents had presented their work and joined the discussion and activities; another five had participated in the discussion and activities without having presented; and five respondents had been ‘silent’ observers of meetings and did not participate in the discussion or activities (for example researchers working on similar topics, who were interested in the discussion, but not involved with the project discussed). Two respondents had more than one role during meetings. Most respondents said that they had previous experience with public engagement. Examples mentioned were participating in the ‘Pint of Science’ festival^35^ ^36^, where researchers talked about their work in an informal pub or café setting, clinical trial engagement, school workshops, art exhibitions, drama, and other related activities like policy development, health workshops or public engagement training.

### Facilitator FGD

All five facilitators, who were three main HREIG group facilitators and two co-facilitators, consented and participated in the FGD. All facilitators are co-authors of this paper, reflecting their contributions in both running the group and conducting the evaluation. Including facilitators as both participants and authors may seem unconventional from a social science point of view. However, as this is an evaluation of a public involvement activity and not a research study, we believe their inclusion was appropriate and necessary. The FGD provided a valuable space for structured reflection, guided by questions from the UK Standards for Public Involvement, and provided important insights for the evaluation.

### Data analysis

FGD transcripts were coded independently by two researchers (AO and SR). Codes were analysed using the Framework Method^37^ and thematic analysis.^38^ Five standards from on the UK Standards framework^32^ were used as initial themes: ‘Inclusive Opportunities’, ‘Working Together’, ‘Support and Learning’, ‘Communications’, and ‘Impact’. We excluded ‘Governance’, as we felt that this standard was more appropriate for an engagement programme rather than a single project. The framework was modified as the data analysis progressed. We combined ‘Support and Learning’ and ‘Impact’ into one theme ‘Learning and Impact’, changed ‘Inclusive Opportunities’ to ‘Supporting diversity and inclusion’ and added it as a sub-theme under ‘Working Together’. We also added the theme ‘Motivations for joining HREIG’ (Table 3). Open-text responses from the HREIG and researcher surveys were also coded and analysed using our modified framework. Likert-scale survey responses were analysed using descriptive statistics.

**Table 3.** Themes and subthemes.

| Theme | Subthemes |
| --- | --- |
| **1. Motivation for joining HREIG** | a. Making a contribution (e.g. to research, society)<br>b. Personal benefits (e.g. learning, professional development) |
| **2. Communications** | a. Perceptions of English as a meeting language<br>b. Communication methods (e.g. for updates, feedback and dissemination of group activities) |
| **3. Working together** | a. Perceptions of meeting format (online and face-to-face)<br>b. Facilitating group discussions<br>c. Supporting diversity and inclusion (recruitment and participant demographics) |
| **4. Learning and impact** | a. Evidence of learning (e.g. skills or personal development)<br>b. Training and support needs<br>c. Impact on MORU research |

## Results

Our framework consisted of four themes, each with two or more subthemes (Table 3): 1) Motivation for joining HREIG, 2) Communications, 3) Working together, and 4) Learning and impact. In the following section, we present the findings from all methods used, according to the themes and their corresponding subthemes.

### Theme 1: Motivation for joining HREIG

#### Making a contribution

Results from both the survey and focus group discussion (FGD) showed that the most common reason for joining HREIG was the desire to make a positive contribution to research and society. Five out of six HREIG members indicated in the survey that this was their primary motivation (Fig. 1). During the FGD participants echoed this view, stating that they wanted to support research that would benefit the wider public. One member explained: *“[an] opportunity to provide feedback or support the researcher. And the research will be shared used for society. Especially, we work based for Thais.*” (FGD, member 4).

**Figure 1:**
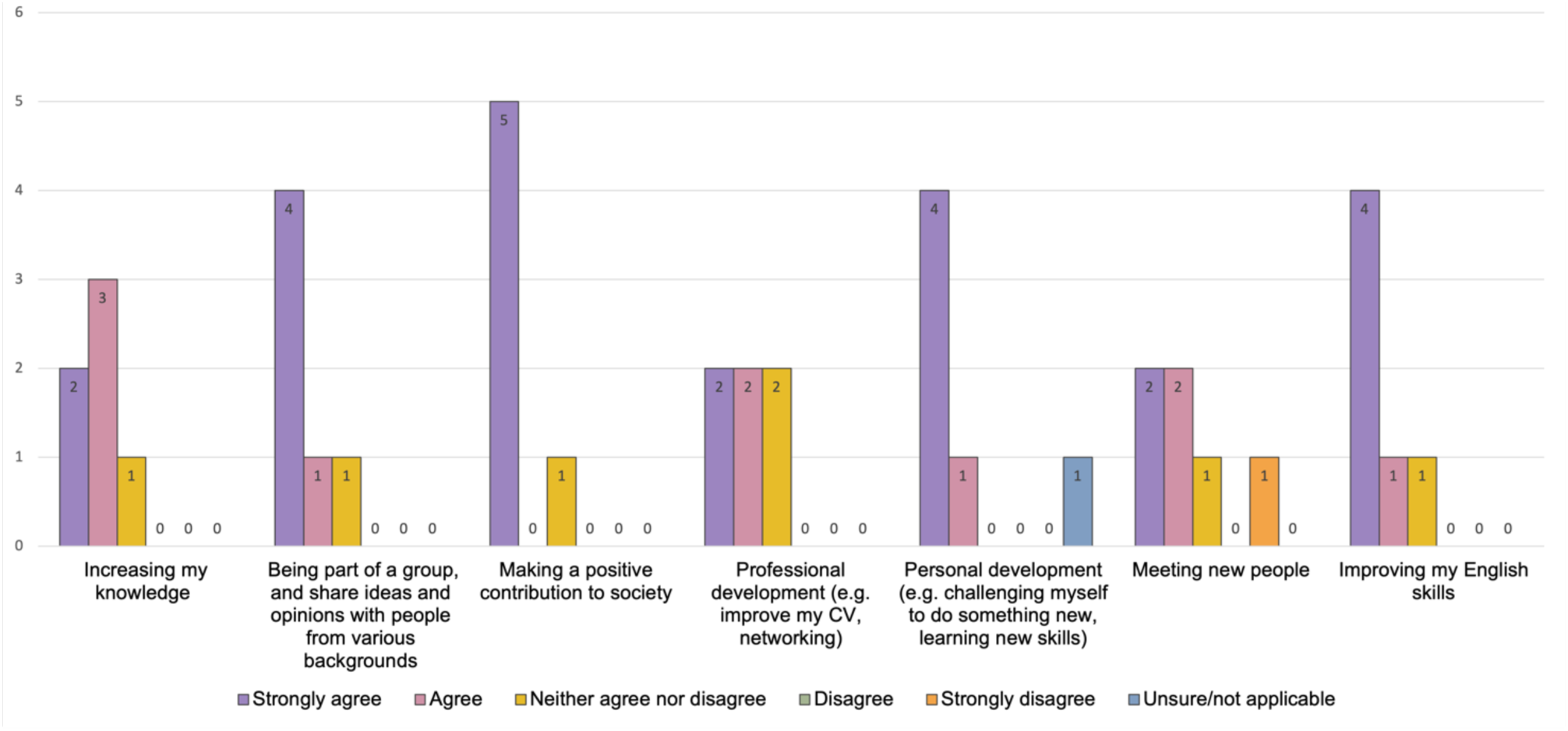
Results from the anonymous group member survey, asking about respondents’ reasons for joining the group.

#### Personal benefits

HREIG members talked about personal benefits as reasons for joining. In the survey and in the FGD, the majority of members said that they wanted to increase their knowledge about science and health research, medicine, and society in general. Most members expressed agreement in the survey that they joined to be part of a group and share ideas and opinions with others, for personal benefits, and to improve their English skills (Fig. 1). Some members indicated in the survey that they joined for their professional development. In the FGD, the younger members expanded on how the group was beneficial as an extracurricular activity alongside their university degree, or their administrative job in a hospital. One member said that they wanted to learn how organisations like pharmaceutical companies or MORU worked. Lastly, some members expressed in the survey and in the FGD that they were interested in trying new things, gaining new experiences and meeting new people:

“*[…] I think it might be like a good idea to get out of my comfort zone and try to do something.”* (FGD, member 2).

In the survey we explored whether members’ expectations about participating in the group had been met (Fig. 2). All members agreed that it had increased their knowledge. Most felt that they were part of a group and able to share their ideas and opinions. Most also agreed that it contributed to their personal development, and that they were able to improve their English skills. When asked if they had been able to make a positive contribution to society, one respondent was unsure, and one disagreed strongly. This suggests a need for clearer communication about group achievements, a view that also was echoed in the member FGD. Half of the members agreed that the group had contributed to their professional development, and half of them said they had been able to network or meet new people. Possibly, the limitations imposed by COVID-restrictions and the move to online meetings affected opportunities for effective networking.

**Figure 2:**
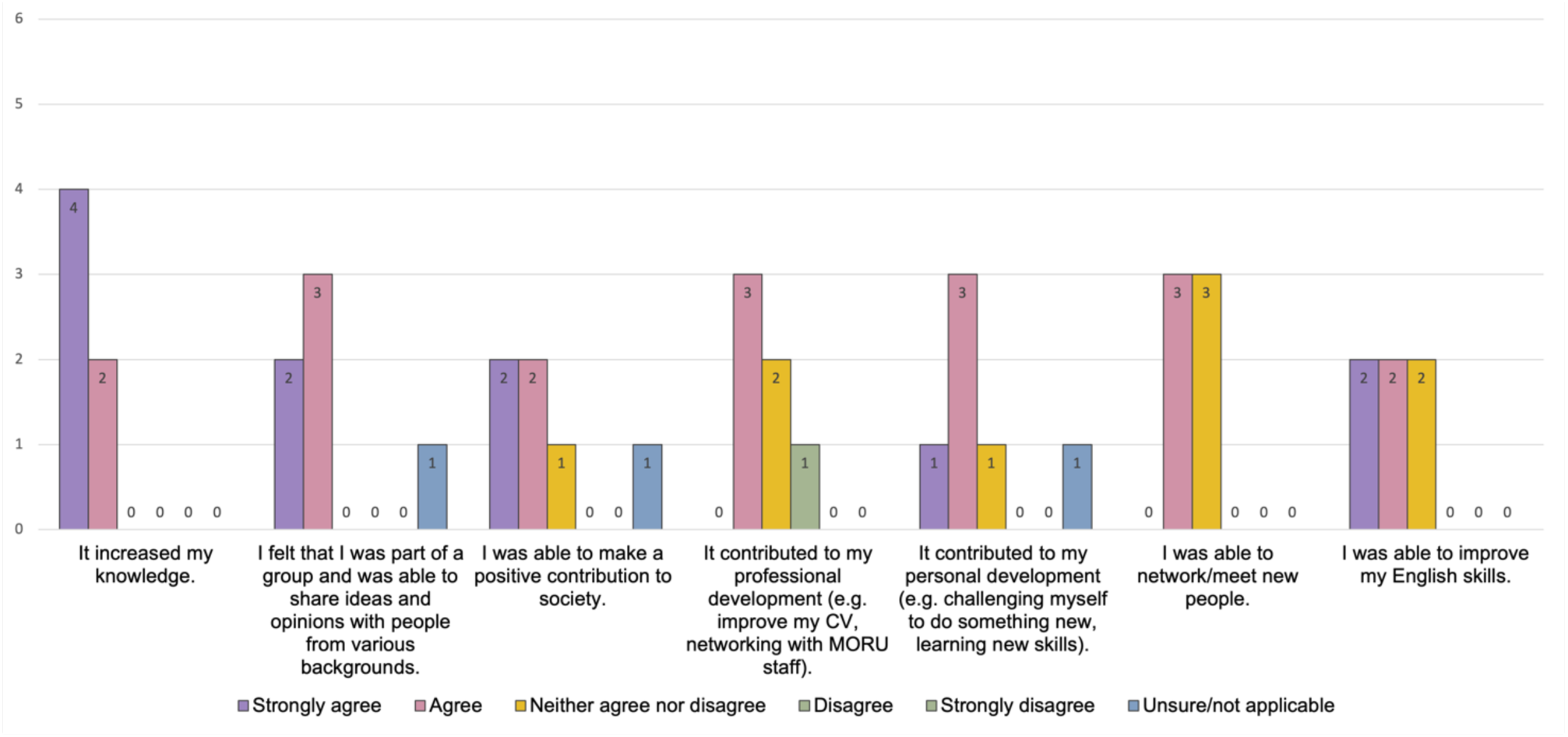
Results from the anonymous group member survey, asking how much respondents agree with the following statements about being an HREIG group member.

### Theme 2: Communications

Guided by the ‘Communications’ Standard in the UK Standards for Public Involvement Framework^32^, we explored the effectiveness of our communications, including the meeting language, and of our processes to feed back to the group and disseminate HREIG’s activities more widely.

#### Perceptions of English as meeting language

Many group members found that English as a main meeting language was challenging, particularly regarding scientific and technical terms (FGD). On the other hand, some members valued the opportunity to practise English in an international environment, and they enjoyed the challenge. Members mentioned the importance of facilitators being at hand to help with translations:

“*[…] in the beginning I think [it was] my language and it was very challenging for me. Sometimes there is some specific vocabulary, and I find it hard, and I feel overwhelmed in the group. But I think [the] facilitator has helped me a lot in every session. I [continued] to stay in the session and become more confident and understand better. I see what in the research, right, I think I’m very proud to be part of this [group].*” [FGD, member 5]

Facilitators expressed concern that the use of English posed a barrier to involvement for some members, who might feel uncomfortable responding, or lose focus during discussions. At the same time, they recognised that using English was a benefit for some members. Facilitators reflected on the need to balance between supporting individual members’ motivation/benefits and equally obtaining everyone’s opinions on a topic. As mentioned earlier, we had switched the main meeting language to Thai in 2022, however, many researchers still presented their work in English. Facilitators acknowledged the importance of their translation support and discussed what could help to research the topic and prepare translations and examples before each meeting, e.g. requesting English presentation slides in advance. In the researcher survey, only one researcher suggested that the meetings should be held in Thai.

#### Communication methods

During the FGD, members expressed a desire to receive updates from researchers about the project’s progress, possibly at a later stage (e.g. one year later). Facilitators (FGD) and researchers (survey) also offered a range of suggestions for how the impact of the group could be communicated better, including project updates at the start of each meeting (a practice already in place), information sharing through internal channels like newsletters and seminars, and externally, e.g. on the community engagement practitioner website Mesh.^39^ The survey also asked researchers for ideas to promote HREIG and the group’s work within and outside of MORU. Their responses included promoting the group in MORU newsletters, emails and the weekly seminar, using word of mouth and social media, and external engagement activities, e.g. with external health or research organisations.

### Theme 3: Working together

We explored the ‘Working Together’ standard^32^ by reflecting on the ways in which we ran our meetings, facilitated group discussion and participation, and how we considered accessibility, inclusion and diversity within the group.

#### Perception of meeting format

Findings from both the FGD and survey indicated that most members preferred meetings in-person. They felt that face-to-face meetings were more interactive and efficient, and they missed physical activities such as grouping ideas on sticky notes. They also said that in-person meetings made communication easier and more natural compared to the online format.

“*Once or twice during online meeting, I was afraid my opinion or the way I talked might upset others. I find it easier to voice my mind when meeting face to face.*” [anonymous member, survey response].

However, they also expressed in the FGD that they felt it was safer to meet online during the COVID-19 pandemic, and that it meant less travel time and no travel expenses. In their opinion, the drawbacks of online meetings were that not everybody was contributing at the same level, which made the discussion less fun, and encountering technical difficulties, e.g. with MS Teams.

Facilitators felt that moving to online meetings made them more accessible for everyone (FGD), which was echoed by members in the ‘LINE’ messenger group chat, particularly as the office location was a bit far for some. Facilitators discussed that they might need to provide more training and technical support, as different levels of online participation require different skills (e.g. more people would have had previous experience using email compared to participating in a Microsoft Teams meeting or using collaborative online tools). They also discussed whether online meetings, which have the option to turn off cameras, made people feel safer to share opinions, but acknowledged that it was easier to raise questions in in-person settings.

#### Facilitating group discussion

The survey responses showed that most members enjoyed being a part of the group (Fig. 3). All members agreed that they felt comfortable to express their honest opinions and interact with the facilitators and researchers, and most members felt comfortable interacting with the others. Everyone agreed that they had been able to make a valuable contribution, and that they felt that the group and MORU worked well together. These responses suggest that members perceived the group as well facilitated, and that they felt comfortable with the setting for discussion. During the FGD, some members expressed that they would enjoy meetings more if participation was higher, which had been a challenge in online meetings.

**Fig. 3:**
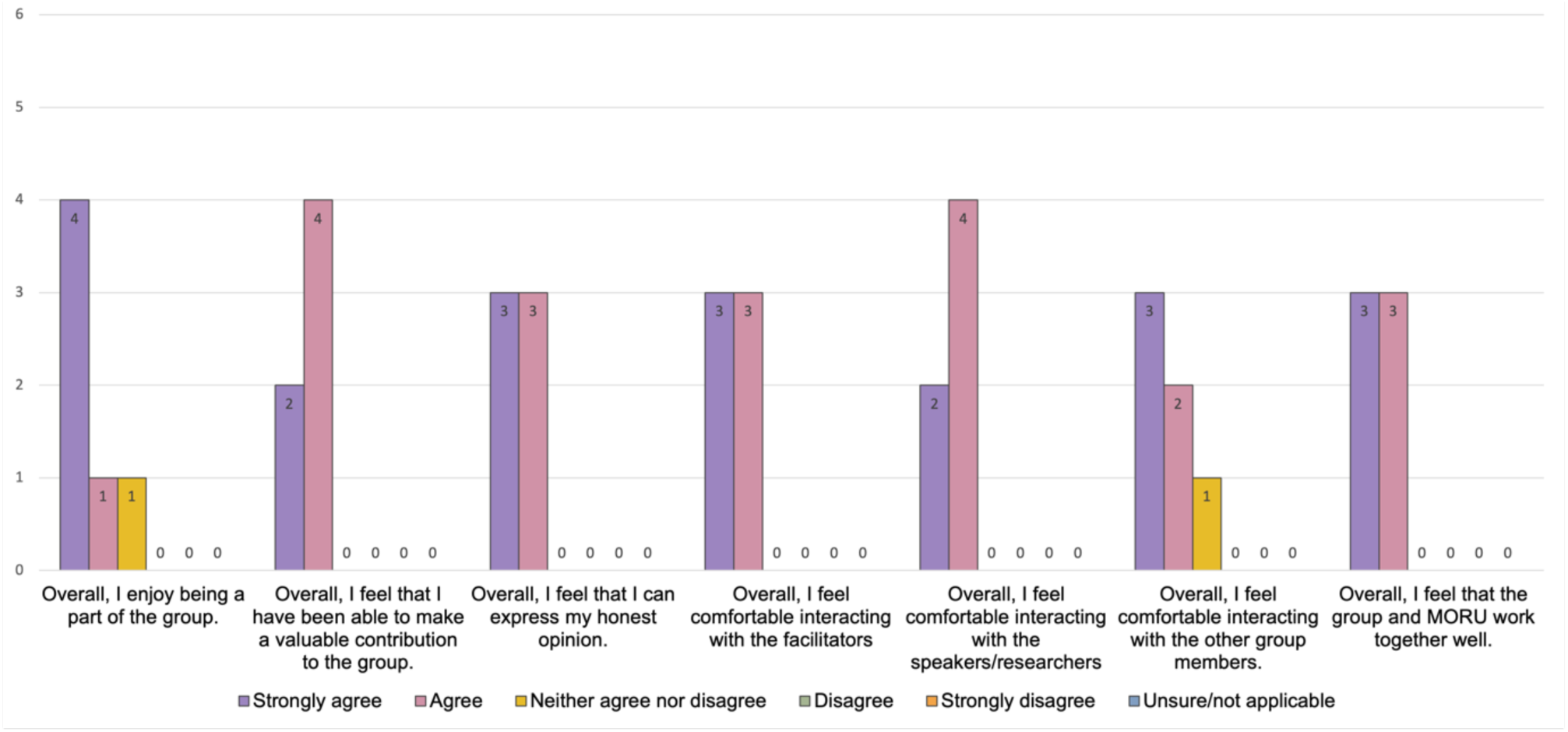
Results from the anonymous group member survey, exploring how respondents felt about being a part of the group.

They suggested using smaller breakout groups of two people for sharing ideas and reporting back to the room, more ice-breaking activities, and using Zoom instead of MS Teams, as they found it easier to use. They also thought that receiving presentation slides in advance would help them to prepare for the discussion.

Facilitators in the FGD reflected on the challenge of getting everyone involved in online discussions, especially participants who were more quiet or reluctant to share their perspectives. The role of the facilitator was seen as very important, not only for moderation and translation, but also for building relationships, which increases trust and encourages members to ask questions without feeling embarrassed. The facilitators said that staffing was resource-intense: HREIG had three dedicated facilitators from the start, and often 1-2 additional staff to help with translations. When reflecting on group participation, facilitator 5 said:

“*And one thing that surprised me, it’s around participants, they are quite open to share and come up with some new things that sometimes you didn’t expect to hear, I don’t know whether it’s a generation or what, but I think they are quite open to speak in in their opinion around the topic that we proposing*.” (FGD, facilitator 5)

Facilitators discussed the importance of recruiting members who felt confident to share their opinions and contribute to the discussion. Lastly, they reflected on the importance of having live translators, as well as the need to send out presentations slides in advance, so that members and translators could get acquainted with the topic beforehand.

#### Supporting diversity and inclusion

Under the ‘Inclusive Opportunities’ Standard^32^, we explored barriers affecting diversity and inclusion, including structural challenges (e.g. meeting location and compensation), communication barriers, and our recruitment process and participant demographics.

In the FGD, members discussed some of the challenges that they experienced, such as not having much background knowledge about medical topics, and people learning at different paces. One member reflected on this in the anonymous survey:

“*I think the challenging part about being a member of HREIG is that you have to learn new information all the time, especially in the field of medicine or public health. Sometimes, it’s not easy to understand all the information in just one session. People have a different pace of learning and understanding. However, I think it is fun to try something out of your comfort zone.”* [anonymous member, survey response].

Some members thought that it would be good to increase participant diversity when recruiting:

“*I felt like we are already doing fine, but it would be great if we could increase the diversity when recruiting, e.g. nationality and occupation, so we can get different types of opinion and more activities to increase teamwork.”* [FGD, member 5]

Some facilitators felt that language was a barrier to involvement (FGD). Even though practicing English was a key motivation for some members, the facilitators worried that others might feel that it was too difficult or too tiring to contribute and remained quiet. Facilitators thought that it was important to have good relationship-building skills to support members with technical or language issues.

Our two recruitment rounds mainly reached professionals, as the advert was posted on a Thai job site. Recruitment also relied on word of mouth through networks of facilitators and members. To widen participants’ profiles, facilitators agreed to be more strategic in the future, revisit recruitment criteria, and seek other avenues to advertise group membership.

In the survey, we asked members about their opinions on compensation. Whilst most thought that the amount was just right, two of members thought it was a bit low. In the FGD, some said that the amount was right if they did not cover travel expenses. Facilitators (FGD) agreed that we needed to compensate participants for their time and felt that the amount was at an appropriate level.

### Theme 4: Learning and impact

We added ‘Impact’ to the ‘Support and Learning’ standard^32^, as we identified learning and development as types of desired impact, in addition to the impact of the group’s feedback on MORU research.

#### Evidence of learning

As described above, all members agreed in the survey that participating had increased their knowledge (Fig 2). In the FGD, members said that they liked experiencing working in an international environment with other members and researchers. They also thought that this was a great way to learn English. Several members mentioned gaining new knowledge about science and society. One member said:

“*[…] I’m not a person who is good at science or pay much attention on science when I was in high school. It did not feel like something relatable for me at all, partly because the curriculum plans. But when I grew up, I think it has become more relatable, especially in the aspect of health. I think these sessions have been very beneficial for me as a person who didn’t know much about [health] science, yeah. So that’s like, I gain more knowledge that is actually useful.”* (FGD, member 3)

They also had gained a better understanding of why we were seeking the opinions of lay people through this group, and how much their perspectives mattered. Members felt proud to be a part of the group and contribute to research. Lastly, they said that they enjoyed getting to know online collaboration tools, which had been new to them.

“*[W]hen my friend told me about the detail of this group, right, I feel like ‘if I’m part of the research, maybe I can help make society better’. But I didn’t really see the full picture until I joined. Then I saw what kind of research you do, how it works, and why our opinions are important to research. I think it’s very exciting to be part of this group.”* (FGD, member 5)

In the survey, researchers responded that the main benefits of engaging with HREIG were hearing the perspectives and opinions of lay people who were not medical researchers; receiving feedback and opinions on their work; and developing their communication skills by practicing dialogue in lay language. One researcher wrote:

“*Getting the lay public view was useful. Helped to focus on the larger picture-how does my project help the general population. Describing my work in lay language was a refreshing challenge*!” Another researcher said: “*I could hear the most unusual type of questions and comments that I would normally be around with. In that sense, it was really eye-opening that I get to know what lay people think about the topics on health and ethics*.” [anonymous researcher, survey response]

Other benefits mentioned were reaching an interested audience for vaccine advocacy and feeling inspiration for engaging similarly in the future. About 2/3 of survey respondents said that their involvement with HREIG helped them to gain more experience in engaging with the public (Fig. 4). Researchers said that they had improved their communication skills, e.g. by discussing science in lay language. They also found it useful to hear opinions from the general public, e.g. for *“[…] addressing concerns that may exist in a study of which we are not aware*”. [anonymous researcher, survey response]

**Figure 4:**
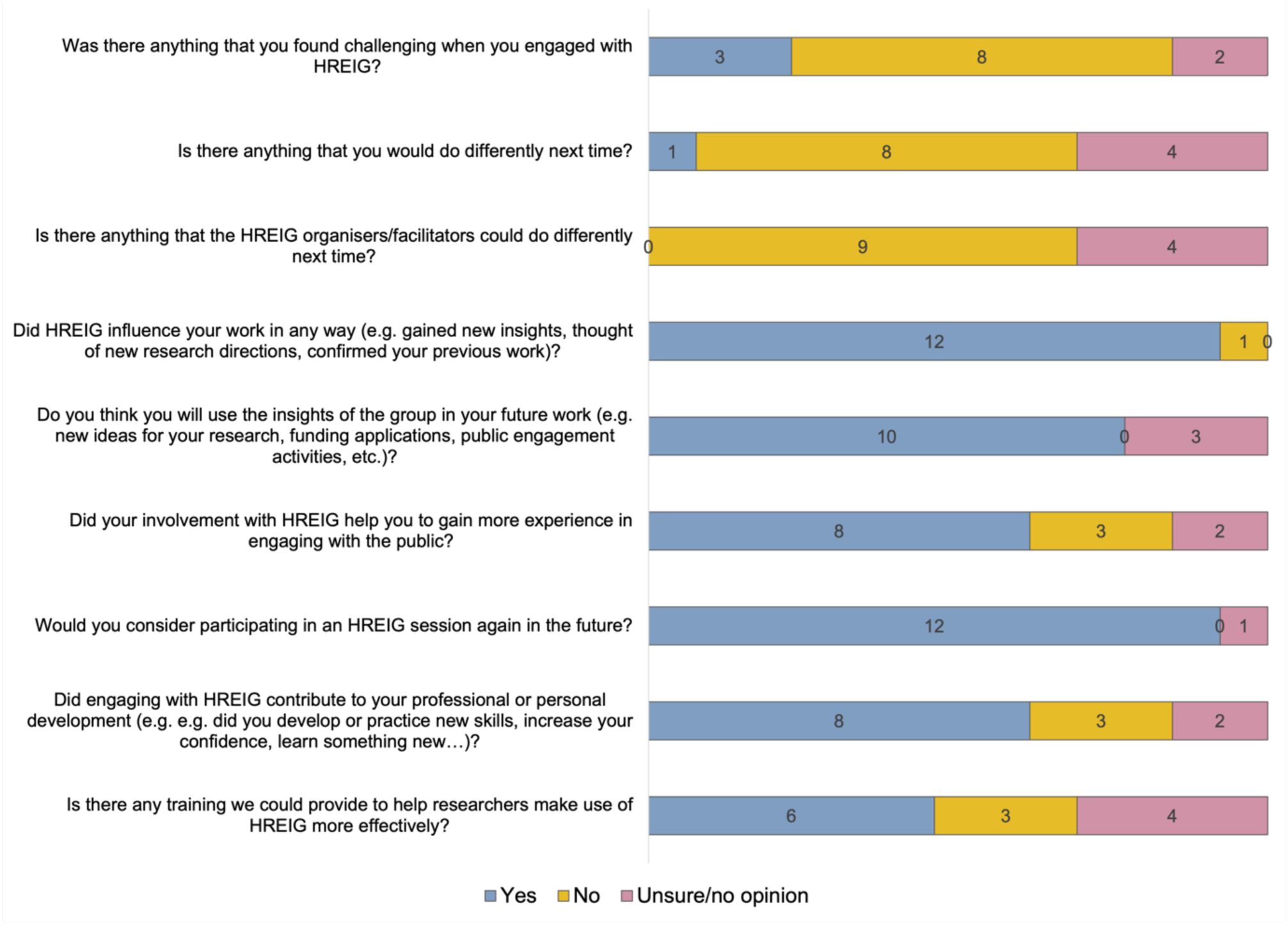
Results from the anonymous researcher survey.

Almost all researchers said that they would consider participating in an HREIG session again in the future (Fig. 4). The reasons given were to gain new insights and experience, both regarding research and public engagement activities, and that engaging with HREIG was a good and useful experience. One researcher who had been an ‘observer’ wrote:

“*If I were to have a new project and want to test out to the public, I will definitely ask HREIG for more help. Even if I am an observer, I still also received many insightful comments that I could apply to my project*.” [anonymous researcher, survey response]

When asked if engaging with HREIG contributed to their professional or personal development, many researchers responded “yes” (Fig. 4). They reported improvements in their public engagement skills, e.g. facilitating discussion and explaining science in lay language. Some also noted it had increased their confidence in presenting their work to a non-researcher audience, developed their social skill, and expanded their network.

Facilitators felt that their skills had improved in many areas (FGD). This included facilitation techniques such as ‘crowd-control’, improvisation, active listening, multi-tasking, constructive group facilitation and supporting people in smaller groups setting and PPI (public and patient involvement). During the COVID-19 pandemic, facilitators gained experience in planning and running online meetings including using different collaborative tools). One non-Thai facilitators remarked that they enjoyed learning about Thai culture and people through discussions with the members. Another facilitator reflected on how they had learnt to trust participants to stay engaged and contribute meaningfully, even when cameras and microphones were turned off. Facilitators also noted that they had learnt how to explain session topics in simple language, and that it often was necessary to start at a more basic level than planned:

“[…] *add value to the research through the help of translation to overcome the barriers of translating scientific stuff into something that they can understand, the group member can understand and provide feedback.*” (FGD, facilitator 4).

By facilitating sessions, they also learnt more about research topics themselves. All facilitators agreed that planning and running meetings was more labour-intensive than thought, but that they all enjoyed it.

#### Training and support needs

When researchers were asked in the survey about challenges in engaging with HREIG, some mentioned hearing answers or opinions that they had not considered from their own researcher viewpoint, and “*to leave space for the discussion and not “intrude” with technical information*”. In response to whether they would do anything differently next time, one researcher suggested conducting the discussion in Thai. There were no suggestions for changes to the meeting organisation and format. This indicates that most researchers had been satisfied with how the sessions had been run. Researchers also suggested ways to engage with HREIG more effectively, such as running practice sessions with mock ‘critique’, training in communication skills to discuss topics in lay language, and storytelling.

Facilitators reflected in the FGD that planning and running the group meetings had been a form of ‘on the job’ training for them. They suggested that there should be more technical training for members, for example provide training on online meetings and collaborative tools for the new members. Facilitators thought that having topic-specific presentations at the start of each session helped place the discussion in context. On the other hand, members who missed a session would also miss this background training, resulting in different levels of understanding among participants. In terms of their own training needs, facilitators suggested in-person and Thai-language training sessions communicating with non-specialist audiences, two-way communication and active listening, improving facilitation skills and preparing facilitators and interpreters with session content in advance.

#### Impact on MORU research

When we asked HREIG members in the FGD about HREIG’s impact on MORU’s research, they highlighted one particular example: their contribution to the development of the “AMR (antimicrobial resistance) Footprint Calculator” website^40^, led by MORU researchers and external partners. Members tested the website before its release and gave feedback on features, such as content and navigation. Their input was incorporated into the final version of the website and members said that they were most proud of this contribution. They also saw their impact as making the research more accessible by helping people from different backgrounds understand the scientific facts or technical terms.

These perceptions were echoed in the researcher survey, where almost all respondents agreed that HREIG had influenced their work in some way. They said that they had gained new insights on their work, particularly regarding the relevance of their research to the public, and that the group’s input had influenced the directions and objectives of their research. One researcher shared that their “*[…] study was facing difficulties with its perception and the use of a particular drug. It was useful to understand from the general public why this was in Thailand […]*”. [anonymous researcher, survey response]

Some said that they now had a better understanding of communication, and how to make language and study concepts less technical and more accessible to non-scientists. They also valued hearing from various backgrounds, although one person pointed out that HREIG may not be able to *“[…] represent the non-responders and all kinds of people in the society*”. [anonymous researcher, survey response]

Almost all researchers indicated that they would use the insights from HREIG in their future work. Some researchers planned to consult lay people before starting future work, and some indicated that they would also like to gain lay opinions on planned public engagement activities or funding applications. One researcher said that they would be “*keen to speak to such a forum before start of future work to align priorities from the researcher’s view with those from the general public*”. [anonymous researcher, survey response]

When asked what the researchers would do to increase HREIG’s impact if they were involved in its organisation, they proposed a range of ideas. Suggestions included providing information or training about the topic beforehand; increasing the number and diversity of members, e.g. including those who might be more critical (e.g. vaccine hesitancy) or who would usually be non-responders; conducting the meeting in Thai; inviting interesting speakers; offering the group some leeway to choose topics for discussion; encouraging face-to-face meetings where possible; and introducing the group and their services to a wider group of stakeholders.

Facilitators saw HREIG as a platform for patient and public involvement (PPI) to provide input into MORU research - particularly at an early stage in the research cycle, before or at the start of projects. They gave some examples (see Table 1), including feedback on the wording of survey questions as part of a MORU-led international COVID-19 study^26^, where the questions were modified for clarification in response to the group’s input. Facilitators emphasised that the group offered a lot of benefits for researchers, particularly in providing them with a different point of view and a lay perspective on their research.

## Discussion

Our findings provide a deeper insight into the motivations and experiences of members of an urban CAB; the opportunities and challenges of running a CAB in English in a non-English speaking country; and the benefits and impact of the group on research and individuals involved, including members, researchers and facilitators.

### Experiences of being an HREIG member

Overall, the feedback of members showed that their expectations for joining HREIG had been met, and that they felt that being an HREIG member brought personal benefits, such as learning about current research or improving their English skills. From the HREIG FGD and survey (Fig. 1-3) two areas stood out in which members’ experiences differed slightly from their expectations: making a positive contribution to society; and networking, meeting new people and feeling comfortable in the group.

Unlike many CABs that focus on certain illnesses or research projects^4^, HREIG was set up to advise on a wide range of studies and topics, including topics of wider societal interest, such as data sharing or artificial intelligence in health care. This could make it more difficult for members to follow how their feedback affected research studies or outcomes. Some projects with more tangible outcomes than others, such as the ‘AMR Footprint Calculator’ website, were easier to follow and therefore more memorable, as evident from the FGD. Members also requested project updates from researchers at a later point.

Being part of a group that is not associated with a research area might also have affected how members felt about connecting with other members. It might be easier to contribute to discussions if everyone has a shared lived experience. The shift to online meetings due to COVID-19 restrictions could have affected the social dynamics and relationships between people who do not know each other well. Bangkok is a ‘transient’ metropolis, as many of its residents move when their circumstances change, e.g. studies or work. This also affected the continuity of the group, e.g. compared to the T-CAB, where members participate in the group for many years.

### Opportunities and challenges of using English as main group language in a non-English speaking country

Our rationale for recruiting members with English proficiency was that our engagement team and the MORU research network are international, with many non-Thai staff and students. We hoped that using English as common language would make it more accessible for researchers and facilitators to talk directly to the group, rather than relying on translators. This was also an experiment, as our other CABs, who serve communities in rural areas, are run in local languages (e.g. Khmer for the Siem Pang youth group^14^). Usually, English proficiency is not common in these areas, and communities might speak several languages or dialects, which creates additional communication challenges.^4^ MORU T-CAB meetings are conducted in multiple languages (Thai/Karen/Burmese/English)^11^ to be as inclusive as possible for its members. However, this makes meetings more resource-intense, as translators and additional time for translating are required during the session and for preparing meeting materials.

Our experience suggests that using English as the common meeting language posed a greater barrier to diversity and inclusion then we had anticipated. While some members enjoyed the opportunity to improve their English, we felt it was harder for all participants to be engaged equally. Even people with good conversational English struggled with scientific or technical terms. It became clear that including English language as recruitment criterion favoured applicants who were mainly students, had a degree, or were professionals working in an international environment. In 2024, Thailand was ranked 106 out of 116 countries, with a slightly higher English proficiency in Bangkok and amongst younger people^41^. Furthermore, a 2018 academic study^42^ reported that higher English utilisation and proficiency in Bangkok businesses was almost exclusively concentrated in downtown areas with mostly foreign companies, hotels, and red-light districts, meaning that including English as recruitment criterion would introduce bias regarding the diversity of group demographics.

### Benefits and impact of the group

When setting up the HREIG, we hoped to see outcomes and impact in two broader areas: benefits for group members, and benefits for MORU (Suppl. Fig. 1). The data from our evaluation study shows that members benefitted personally, by learning about new topics and practicing language and digital skills. They also found enjoyment in being a part of the group and contributing to research. This resonates with feedback from other CABs, e.g. members of the T-CAB at the Thai-Myanmar border described the CAB as a ‘place to learn’.^11^ The benefits for MORU of running the HREIG were evident in the impact it had on the researchers and facilitators that participated, and in the changes to MORU research and engagement projects. Researchers clearly valued the opportunity to get in touch with members of the public who are affected by their research.

The Bangkok HREIG is part of a larger network of MORU CABs. There are differences in the communities that they serve, and this is reflected in the kind of impact that the groups’ feedback has on MORU research. For example, members of the Tak Province CAB at the Thai-Myanmar border (T-CAB)^11^ ^12^ consider the demographics of the communities living at the border (e.g. migrants and refugees, ethnic groups, pregnant women and babies), and the unique barriers they face when accessing clinical trials and health services (e.g. having no documentation, traveling during rainy season). A lot of T-CAB’s impact is highlighting how research study protocols should be modified to consider vulnerable groups, and to mitigate burdens faced by patients and participants. The Chiang Rai hill tribe CAB (CR-CAB)^9^ works with hill tribe ethnic groups who mostly live in rural and remote areas and have no written language forms. As this is a significant barrier to accessing health services and participating in research, the CR-CAB provides a lot of advice on communication and translation issues, e.g. how to communicate research concepts to communities who do not have any previous experiences of research, and how to make informed consent forms and participant information materials used in clinical studies at the Chiang Rai site more accessible^9^.

The Bangkok HREIG complements this work by advising on topics of broader societal interest. For example, they suggested how to make the wording of key messages used in Thai press releases clearer to lay people, and offered their opinions on whether researchers should be able to re-use de-identified data from clinical trials in other studies without obtaining informed consent again. The HREIG was the only CAB in our network that was able to continue meeting during COVID-19 restrictions, due to their ability to reliably access mobile devices and the internet. The group even met more frequently during the early pandemic to advise on ongoing COVID-19 studies (see Table 1). HREIG members were also able to test digital engagement activities, e.g. they provided feedback on collaborative online activities that we had developed for the ‘AMR Dialogues’ public engagement project^43^, and they tested the ‘AMR Footprint Calculator’^29^ website before its release. As the website project was very tangible, members mentioned it most frequently when asked about the impact of the group. Due to their English proficiency, some members attended meetings at MORU as a public representative and independent member, e.g. the in-person MORU Governing Board meeting, or joined steering committees for clinical trials.

Overall, the work of our CABs complements each other in impact that they have on MORU research. It would not be feasible to set up one MORU CAB that ‘represents’ all or most the groups who are involved in or affected by our research – this diversity is achieved through our CAB network. In addition, our CAB network is not the only source of community input, and other kinds of impact are realised by other forms of engagement.

### Strengths and limitations

As this study was an evaluation, the number of participants was small. However, even though this means that our findings are not generalisable, they were important for our context and resulted in changes made to the running of the group. We also hope that they will be of interest to those running advisory groups or looking to establish advisory groups in similar settings. Reflecting on the methods, we chose focus group discussion as a method, as it allows participants to interact and build on each other’s ideas^44^, and it is less resource-intense than individual in-depth interviews. However, semi-structured individual interviews can yield more detailed responses. The FGD facilitators were also the HREIG facilitators, rather than impartial external evaluators, and this could have affected honest or critical feedback. To mitigate this, we included the short anonymous survey at the start of the member FGD, and its responses were similar to the discussion.

### Lessons learnt

Our findings have informed the development and recruitment of members for ‘HREIG 2.0’, following the conclusion of the first group’s term. We expanded the group size to 20 members to increase diversity of the group and mitigate the impact of individual absences. Meetings are now held monthly and in-person to keep members engaged. At the start of each session, we provide session recaps and updates and share feedback from the researcher after they consulted the group. Sessions now operate in Thai, and if the presenter does not speak Thai, we offer translation and assistance during the session We also ensure that the use of jargon is minimised.

### Conclusions

The findings of this evaluation study offer an in-depth insight into the motivations and experiences of group members who participated in the HREIG community advisory board, and the experiences of facilitators and researchers who engaged with the group in that period. There are only few publications describing the running and evaluation of public advisory groups who operate in an international setting, where group members, facilitators and researchers speak different languages. We hope that our findings will contribute to the knowledge in this field and help others who would like to set up a community advisory board in a similar context.

## List of abbreviations

FGD: focus group discussion

HREIG: Health Research and Ethics Interest Group

MORU: Mahidol-Oxford Tropical Medicine Research Unit

## Declarations

### Ethics approval and consent to participate

Ethics approval was granted by the Oxford Tropical Research Ethics Committee (OxTREC, reference 555-21), and the Ethics Committee of the Institute for the Development of Human Research Protections (IHRP, reference IHRP2021161). Informed written consent was obtained from all participants.

### Consent for publication

Not applicable.

### Availability of data and materials

Data underlying the paper may be requested from the Mahidol-Oxford Tropical Medicine Data Access Committee.

## Authors’ contributions

PYC led the establishment of the HREIG. AO, SR and PYC coordinated recruitment and group activities from 2019-2023. All authors participated in facilitation of the group activities. AO led the design of the evaluation study and the data collection, supported by the other authors. SR, TP and BN supported translation and submission of the ethics approval documents in Thailand. AO and SR analysed the data, guided by BN. AO wrote the first draft of the manuscript. All authors contributed to the writing of the manuscript and reviewed and approved the final version of the manuscript. AO is the principal investigator and the guarantor of the paper.

## Acknowledgements

We would like to thank the HREIG members for their valuable contributions over the last years, and for sharing their experiences in our evaluation study. We also thank the researchers who participated in our survey and provided useful feedback. Lastly, we thank Napat Khirikoekkong for critical reading of the manuscript.

