## Supplemental Figure 1 for "Evaluation of the Bangkok Health Research and Ethics Interest Group: reflecting on the experiences of group members, researchers and facilitators participating in an urban community advisory board in Thailand"

**Supplementary Figure 1:** HREIG logic model.

### INPUTS

**People and organisations:**
- MORU Bioethics and Engagement staff
- Researchers
- Members of the public

**Skills and knowledge:**
-PE Facilitators
-Researchers

**Resources**:
- Time (i.e. planning and running of group sessions, debrief)
- Money (i.e. Wellcome grant, tokens of appreciation, refreshments)
- Spaces at Mahidol University

### Activities

Member recruitment

Scheduling and organisation of meetings

Planning of evaluation

---

Group meetings and discussions

Training in bioethics and research principles

Icebreakers and socialising

Facilitation and minute-taking

Mentoring researchers in making their research accessible

Communication to group members

---

Debrief sessions for staff and researchers

Reflection, monitoring and formal evaluation

Reporting to researchers, the group and funding bodies

### OUTPUTS

Group member number, demographics and meeting attendance

Number of researchers and external partners involved

Number of meetings and session agendas

A set of interactive activities

Information materials

Logic model

'Pint of Science' attendance

---

Number of projects and topics discussed

Meeting minutes

Recommendations and suggestions for MORU research and other activities

Session feedback by members and researchers

---

'Lessons learnt'

Reports (i.e. evaluation) and case studies

### OUTCOMES

#### Benefits for the group

**Short-term (0-12 months)**
Group members gain knowledge and understanding about health research and bioethics.

Members understand how MORU programmes run and what the ethical issues are.

Members enjoy being a part of the group.

Members feel that their opinion and expertise are valued.

Members feel that they benefit from participating.

**Medium-term (12-24 months)**

Members have sufficient knowledge to provide honest and critical opinions on complex issues.

Members feel confident in communicating with and advising researchers.

MORU staff and group members work together effectively as equal partners.

The group has a high profile within the MORU network.

#### Benefits for MORU

**Short-term (0-12 months)**

Researchers develop or practice skills in talking to a lay audience.

Researchers feel that they benefited from interacting with the group.

Researchers use the inputs received to make improvements to their programme, patient communication or other areas.

MORU engagement staff develop their group facilitation skills.

**Medium-term (12-24 months)**

Researchers proactively seek out the group's advice in the early stages of the research cycle.

The Department for Bioethics and Engagement gains experience in working with a public group, and a better understanding of the opportunities and limitations of this approach.

### IMPACT

MORU programmes use the views of the HREIG to make their protocols and communications more ethical.

The partnership between the group and MORU staff and researchers helps to build trust in MORU programmes.

Being a part of the group makes group members feel knowledgeable and empowered.

The group is an integral part of the MORU research life cycle.

The lessons learnt form the project inform similar group setups within and outside of MORU.

*Based on the 'Logic Model Development Guide' by the W.K. Kellogg Foundation (2004)*
